# Epidemiological identification of a novel infectious disease in real time: Analysis of the atypical pneumonia outbreak in Wuhan, China, 2019-20

**DOI:** 10.1101/2020.01.26.20018887

**Authors:** Sung-mok Jung, Ryo Kinoshita, Robin N. Thompson, Katsuma Hayashi, Natalie M. Linton, Yichi Yang, Andrei R. Akhmetzhanov, Hiroshi Nishiura

## Abstract

**Objective:** Virological tests indicate that a novel coronavirus is the most likely explanation for the 2019-20 pneumonia outbreak in Wuhan, China. We demonstrate that non-virological descriptive characteristics could have determined that the outbreak is caused by a novel pathogen in advance of virological testing.

**Methods:** Characteristics of the ongoing outbreak were collected in real time from two medical social media sites. These were compared against characteristics of ten existing pathogens that can induce atypical pneumonia. The probability that the current outbreak is due to “Disease X” (i.e., previously unknown etiology) as opposed to one of the known pathogens was inferred, and this estimate was updated as the outbreak continued.

**Results:** The probability that Disease X is driving the outbreak was assessed as over 32% on 31 December 2019, one week before virus identification. After some specific pathogens were ruled out by laboratory tests on 5 Jan 2020, the inferred probability of Disease X was over 59%.

**Conclusions:** We showed quantitatively that the emerging outbreak of atypical pneumonia cases is consistent with causation by a novel pathogen. The proposed approach, that uses only routinely-observed non-virological data, can aid ongoing risk assessments even before virological test results become available.

## INTRODUCTION

A cluster of cases of atypical pneumonia with unknown etiology in Wuhan, China attracted global attention at the end of 2019 (Wuhan Municipal Health Commission, China, 2019; World Health Organization, 2020). An impressive series of rapid virological examinations ruled out common pneumonia-causing viruses such as influenza viruses, adenoviruses, and the coronaviruses associated with Middle East respiratory syndrome (MERS) and severe acute respiratory syndrome (SARS) (Wuhan Municipal Health Commission, China, 2019; Normile, 2020a, 2020b; World Health Organization, 2020). As of 12 January 2020, the causative agent is suspected to be a coronavirus of non-human origin (European Centre for Disease Control and Prevention, 2020; Normile, 2020b).

While examination of the viral genome is critical for identifying the pathogen, information made publicly available in real time describing clinical characteristics and other outbreak-related factors can also allow experts to consider the etiology and thereby differential diagnoses. For instance, most cases shared a history of visiting or working at a seafood market in Wuhan (Wuhan Municipal Health Commission, China, 2020), where exposure to the novel coronavirus is suspected to have occurred with no evidence of direct human-to-human transmission (World Health Organization, 2020), leading us to believe that the cluster of cases was due to “Disease X” (i.e., an infectious disease of previously unknown viral etiology). However, rigorous quantitative assessment of the chance that the disease is in fact Disease X has not previously been undertaken. The present study addresses this, demonstrating that non-virological information can lead to an objective classification of Disease X, using a simple statistical model that exploits the well-known Bayes’ theorem.

## METHODS

As the outbreak unfolded, we calculated in real-time the probability that the pathogen responsible for the atypical pneumonia was novel (Disease X), or whether instead the outbreak was generated by a previously known pathogen that can cause pneumonia. Our analysis began on 30 December 2019, when the Wuhan Municipal Health Commission announced that there had been a surprisingly large number of atypical pneumonia cases. At that time, we assumed the causative agent could have been one of seven known viral or three known bacterial diseases, along with the chance that it was instead Disease X. We tracked two of the most active medical social media sites, i.e., ProMED (ProMED, 2020) and Flutracker (Flutracker, 2020), that reported the non-virological characteristics of the outbreak, including atypical pneumonia, other clinical characteristics, and exposure factors, as it progressed. These characteristics do not necessarily represent the features that were causing disease, but are instead basic observations from the ongoing outbreak. Given these characteristics, we then calculated the probability that the ongoing outbreak is due to a known disease or unknown Disease X. On the first day of calculation (i.e. 30 December 2019), the only explanatory factors we included was atypical pneumonia, which was common to all enumerated diseases. Our analysis represents simple logical deductions from the limited data that were available during the outbreak in a quantitative manner and was updated to reflect new information about the outbreak as it became available in real time.

Table 1 shows the information compiled about the current outbreak, and the dates on which each of these characteristics were discovered. Each characteristic listed was assigned a value of zero or one, denoting whether or not the characteristic of listed outbreak, not individual cases, was likely for the emerging outbreak, and the equivalent values for outbreaks of previously observed pathogens were also noted. We make two assumptions to use and un-use a part of the input exposure characteristics: (i) previously known disease outbreaks are all based on empirically observed notion (and do not include the new exposure data (i.e., exposure at a wet market), that is specific to novel coronavirus in Wuhan, which may be non-informative to other outbreaks for the calculation) and (ii) all exposure characteristics are known for all previously known outbreaks, incorporating all factors enumerated. Also, once pathogens were ruled out as the causative agent of the current outbreak, they were removed from our analysis: for example, highly pathogenic avian influenza (HPAI) (H5N1) was confirmed not to be the causative agent by laboratory testing on 3 January. Hence, we omitted this pathogen from our analysis from 3 January 2020 onwards.

**Table 1.** Characteristics of outbreaks driven by pneumonia-causing pathogens, with respect to the current outbreak in Wuhan, China.

| Category | Characteristic | Current outbreak |  | Viral outbreaks |  |  |  |  |  |  | Bacterial outbreaks |  |  |
| --- | --- | --- | --- | --- | --- | --- | --- | --- | --- | --- | --- | --- | --- |
|  |  | Disease X | Date | SARS* | MERS** | HPAI***<br>(H5N1) | HPAI***<br>(H7N9) | Other<br>influenza<br>viruses | Adenoviruses | Hantaviruses | *Chlamydia*<br>*pneumoniae* | *Mycoplasma*<br>*pneumoniae* | Legionellosis |
|  |  |  | info<br>shared |  |  |  |  |  |  |  |  |  |  |
| Clinical | Atypical pneumonia | 1 | 30-Dec | 1 | 1 | 1 | 1 | 1 | 1 | 1 | 1 | 1 | 1 |
| Clinical | CT (pulmonary infiltrates) | 1 | 31-Dec | 1 | 1 | 1 | 1 | 0 | 0 | 1 | 1 | 1 | 1 |
| Clinical | Low white blood cell counts | 1 | 31-Dec | 1 | 1 | 1 | 1 | 1 | 1 | 1 | 0 | 0 | 0 |
| Clinical | No response to antibiotics | 1 | 31-Dec | 1 | 1 | 1 | 1 | 1 | 1 | 1 | 0 | 0 | 0 |
| Clinical | Frequent human transmission | 0 | 31-Dec | 1 | 1 | 0 | 0 | 1 | 1 | 0 | 1 | 1 | 1 |
| Clinical | Substantial lethal cases | 0 | 31-Dec | 1 | 1 | 1 | 1 | 0 | 0 | 1 | 0 | 0 | 0 |
| Travel/Occupation | Worked/visited a wet market | 1 | 31-Dec | 0 | 0 | 1 | 1 | 0 | 0 | 0 | 0 | 0 | 0 |
| Travel/Occupation | Worked/visited a hospital | 0 | 31-Dec | 1 | 1 | 0 | 0 | 0 | 0 | 0 | 0 | 0 | 0 |
| Travel/Occupation | Visited Middle East countries | 0 | 31-Dec | 0 | 1 | 0 | 0 | 0 | 0 | 0 | 0 | 0 | 0 |
| Travel/Occupation | Visited hot spring or contact<br>with potable water | 0 | 31-Dec | 0 | 0 | 0 | 0 | 0 | 0 | 0 | 0 | 0 | 1 |
| Zoonotic | Contact with camels | 0 | 31-Dec | 0 | 1 | 0 | 0 | 0 | 0 | 0 | 0 | 0 | 0 |
| Zoonotic | Contact with parrots/wild birds | 0 | 31-Dec | 0 | 0 | 1 | 0 | 0 | 0 | 0 | 1 | 0 | 0 |
| Zoonotic | Contact with rodents | 0 | 31-Dec | 0 | 0 | 0 | 0 | 0 | 0 | 1 | 0 | 0 | 0 |

\*Severe acute respiratory syndrome; \*\*Middle East respiratory syndrome; \*\*\*Highly pathogenic avian influenza. Zeros represent characteristics that are unlikely for outbreaks for that pathogen, and ones represent characteristics that occur. Dates and characteristics for the ongoing outbreak were obtained from two online information systems [5,6], and information for other pathogens was summarised from the pathogen-specific pages on the WHO and CDC websites.

To assess the probability that the emerging outbreak was caused by a variant of a known pathogen, we first calculated the distance between the set of characteristics of the ongoing outbreak and those of previously known pathogens. The distance between the characteristics of the ongoing outbreak and cases due to pathogen *j* is denoted by *d*_*j*_. We assumed that the probability that the outbreak is due to a variant of pathogen *j* decreases exponentially with distance *d*_j_. Then, by Bayes’ theorem,

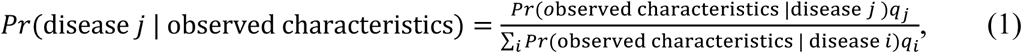

in which the sum in the denominator is over all possible diseases *i* (i.e. each of the columns of Table 1, including the column describing the current outbreak). The constants *q*_i_ are *a priori* probabilities that the outbreak is due to pathogen *i* (Nishiura et al., 2012; Ejima et al., 2014). We set uninformative priors for all pathogens considered, so that *q*_*i*_ was simply the reciprocal of the number of pathogens being considered (including Disease X) on each date in our analysis. We initially estimated the distance between observed characteristics of the outbreak and each known candidate pathogen using the Hamming distance (i.e., the sum of squares differences between the entries in the columns of Table 1 corresponding to the Disease X and the candidate pathogen). Then, we assumed that the probability that the outbreak is driven by disease *j* is governed by a negative exponential function

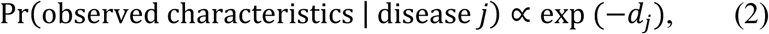

where *d*_j_ is the calculated Hamming distance.

We also repeated our analysis using an alternative measure of the distance between observed characteristics of the outbreak and each known candidate pathogen, namely the Euclidean distance (i.e. the square root of the Hamming distance). In each case, we assumed that the importance of each characteristic had an identical weight in our analysis, so that a simple quantitative assessment could be obtained in a probabilistic manner without the need for subjective judgement.

Combining equations (1) and (2), and assuming that *q*_i_ is identical over *i*, we have:

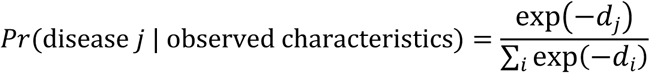

The probability that the outbreak is driven by Disease X corresponds to the distance *d*_X_= 0, and represents a risk score taking values between the reciprocal of the number of candidate pathogens including Disease X itself and one:

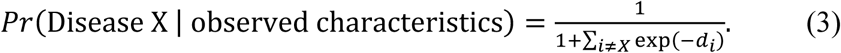

Supposing that there are *n* known pathogens responsible for the atypical pneumonia, the probability of observing Disease X without any information is identical with the probability of observing other listed pathogen (i.e., 1/(1+*n*)) and as pathogens are ruled out by laboratory testing, the identical probability increases (i.e., 1/11 until 2 Jan 2020, 1/7 from 3 Jan 2020 and 1/5 from 5 Jan 2020 in current outbreak). In addition, if the probability of observing Disease X according to equation (3) takes a value close to the probability of observing other candidate pathogens, the overall probability that the outbreak is due to a novel pathogen should be interpreted as being low. A result of significant practical importance, however, is when the probability of observing Disease X is close to one or much larger than the probability corresponding to each previously observed candidate pathogen. In that case, all candidate pathogens are not similar to the causative agent of the ongoing outbreak, and so the outbreak is likely to be due to a novel pathogen.

We converted the probability of disease X into the equivalent percentage value (so that, for example, a result of 0.8 in equation (1) is assumed to mean an 80% probability) and refer to the percentage value as the “probability of Disease X” hereafter.

## RESULTS

We show temporal changes in estimates of the probability that the ongoing outbreak is driven by each candidate pathogen in Figure 1. Because the only information on 30 December 2019 was that cases displayed symptoms of pneumonia, the distance between ongoing outbreak and known ten diseases was all zero, and thus, all eleven candidate pathogens initially showed an identical probability of 9.1% (i.e., 1/11). Additional characteristics became known the following day (i.e., 31 December 2019), and consequently, the inferred probability that the outbreak was driven by a novel pathogen increased substantially to 58.6% and 36.9% for Hamming and Euclidean distance metrics, respectively. When the exposure characteristic (i.e. exposure at a wet market), that is specific to ongoing outbreak were excluded from the analyses, the probability of observing Disease X given observed characteristics is as high as 48.7% and 32.6% for Hamming and Euclidean distance.

**Figure 1.**
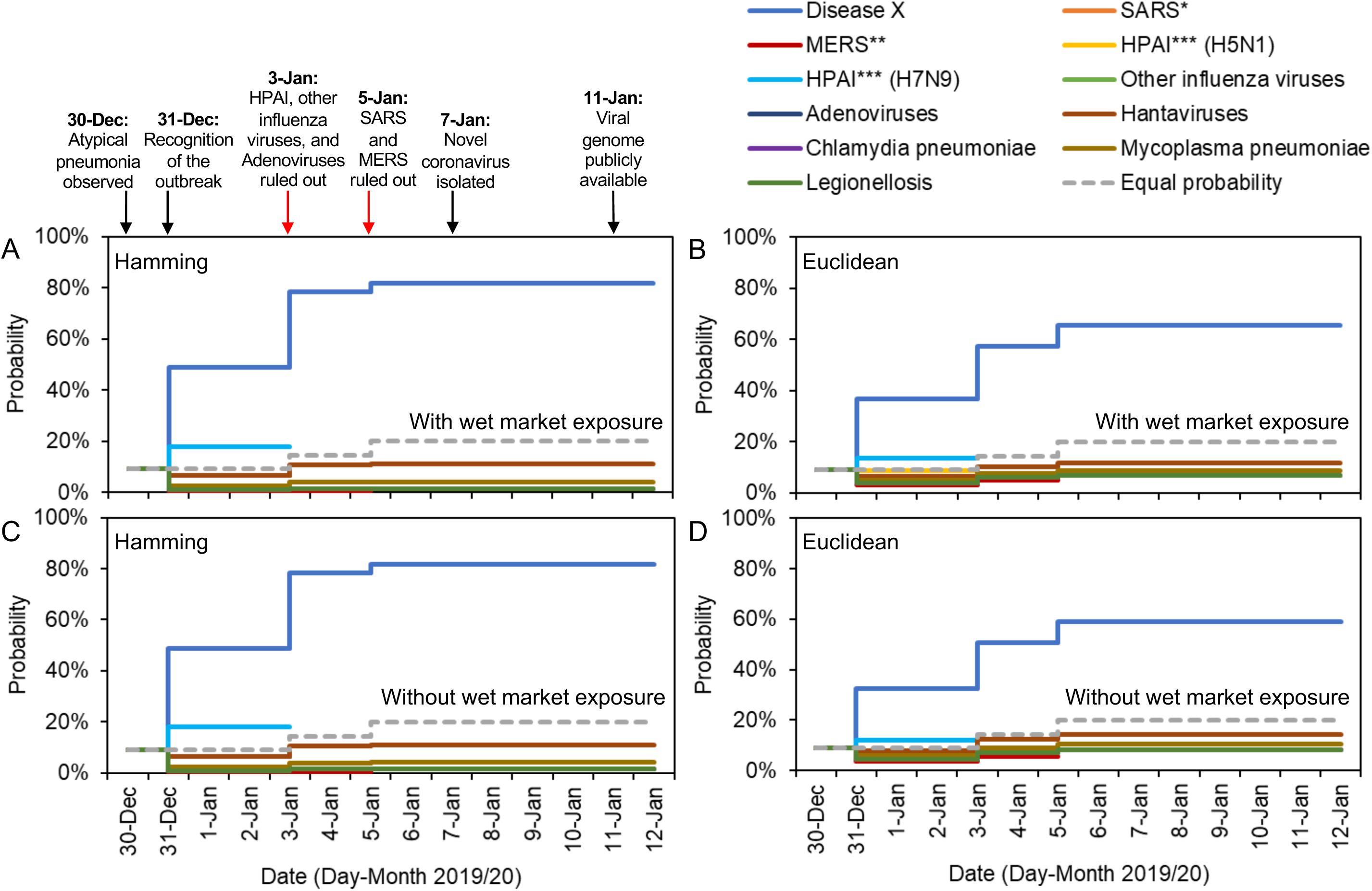
Real-time estimation of the probability that the ongoing pneumonia outbreak is driven by each candidate pathogen, given available information at different timepoints. The probability that the outbreak is due to an unknown pathogen (Disease X) increases as more information becomes available, since the unknown pathogen can be seen to exhibit characteristics dissimilar to those observed in previous outbreaks, and since known pathogens are ruled out by laboratory results. Arrows indicate new information available on each date. Results are shown for different metrics describing the distance between characteristics of the ongoing outbreak and each candidate pathogen, and by knowledge (inclusion or exclusion) of exposure characteristics of Disease X (i.e. Work/visited a wet market), specifically: A. Hamming distance (the sum of squares difference between the entries in the columns of Table 1 corresponding to the ongoing outbreak and the candidate pathogen considered) with wet market exposure; B. Euclidean distance (the square root of the Hamming distance) with wet market exposure; C Hamming distance without wet market exposure; D Euclidean distance without wet market exposure. Dashed grey line shows the probability without considering any information except atypical pneumonia (i.e. equal probability=1/(1+number of candidate pathogens)). Note that the probability of some diseases is identical, for example, SARS and *Mycoplasma pneumoniae* has equal probability from 30 Dec to 4 Jan, and Legionellosis and *Chlamydia pneumoniae* has equal probability from 30 Dec to 12 Jan (Details in Supplementary material 1).

Later in the outbreak, adenoviruses, HPAI (H5N1 and H7N9) and other influenza viruses were ruled out on 3 January 2020, leading the probability of Disease X being assessed as 90.7% and 57.2% for Hamming and Euclidean distance metrics, when all factors were considered as characteristic. Excluding the wet market exposure, the probability of Disease X was 78.2% and 50.6% for Hamming and Euclidean distance metrics, respectively. SARS- and MERS-associated coronaviruses were ruled out as the causative agent on 5 January 2020, leading to a very high estimate for the probability that the outbreak is caused by a novel pathogen once all information had been collected. As of 12 January 2020, the probability of Disease X is estimated to be 92.5% and 65.5% using the model considering all the factors, while the model excluding the characteristic of exposure at the wet market indicated that the probability of Disease X is assessed as 81.8% and 59.1% for Hamming and Euclidean distance models, respectively

## DISCUSSION

In this analysis, we have shown quantitatively that the ongoing outbreak of pneumonia cases in Wuhan has almost certainly been caused by a novel pathogen. This was demonstrated using a series of clinical, occupational, and behavioral observations extracted from fragmented reports describing the cases as these reports became available in real time (European Centre for Disease Control and Prevention, 2020; Wuhan Municipal Health Commission, China, 2020). Although virological investigation is the gold standard for pathogen identification and a novel coronavirus has now been identified from some of the cases, such laboratory-based outcomes can only be obtained after successfully sequencing the novel virus, which can be a lengthy process. It still remains for the microbiological causal link to be established, for instance by ensuring that Koch’s postulates are met (e.g., as seen in a study of Zika virus (Krauer et al., 2017)). In the ongoing outbreak, the provisional identification of a novel coronavirus was performed on 7 January 2020 and announced formally on 9 January 2020 (World Health Organization, 2020). We have shown that non-virological information can indicate that the cause of the outbreak is likely to be a novel pathogen, and that this conclusion could have been obtained before virological test results were announced. Disease X was inferred to be very likely on all dates from 31 December 2019 onwards—the date on which descriptions of outbreak characteristics began to emerge.

When sufficient clinical details of cases (e.g., complete blood cell counts) are available, the number of causative pathogens considered can be limited to a reasonable number. In this instance, atypical pneumonia combined with reduced white blood cell counts and the lack of response to antibiotics indicated that the pathogen was consistent with viral rather than bacterial infection. With such information, collecting non-virological data can lead to a convenient quantification of the probability of Disease X, while awaiting the results of virological tests. We believe that the proposed approach can greatly improve the ongoing risk assessment practices across the world.

It is critically important to discuss two issues that the definition of variables in Table 1 has involved. First, a critical underlying assumption is that Table 1 reasonably represents outbreak characteristics of ongoing and previously known outbreaks. The representation does not reflect observation from all confirmed cases nor epidemiological findings from a case control study (e.g. statistically significant risk factor). Rather, zeros and ones in the table were defined in a phenomenological manner. Depending on readers, the defined nominal values can be different from what it was shown in Table 1 and ours is only for the exposition using a typical Table 1 that authors came up. Second, as we have shown, there are multiple combinations of characteristic data to be used. Namely, as an exposure to a wet market for known disease outbreaks other than HPAI was not necessarily derived from empirical observation, the fairness of an assumption that the majority of cases of those known disease outbreaks were asked not to have visited a wet market would be a subject for debate.

In the past, descriptive outbreak information has been used to produce sensitive outbreak case definitions, and causative agents have been pinpointed without using statistical methods in combination with epidemiological observations. In the present study, we have shown that such assessments can be made quantitatively using a simple statistical model, allowing for comparison of the likelihood of causative agents among all possible candidates. When outbreak characteristics are shared and updated in real-time (Table 1), these data can contribute to narrow down the possible range of causative agents. In the case of the outbreak in Wuhan, our calculation of the probability that each pathogen is the causative agent indicates that virologically excluding the possibility of influenza viruses, adenoviruses and known virulent coronaviruses associated with SARS and MERS on 3 and 5 January 2020 can be regarded as an “unsurprising” finding.

As important limitations, the precision and credibility of input data, and the method for calculating the distance between candidate diseases and the observed outbreak, must be refined in the future. First, our proposed approach used very limited data in Table 1 for logical quantification of the probability that each pathogen was the causative agent. However, with more clinical data, the dataset of characteristics could be replaced by continuous frequencies (e.g. the frequencies of cases experience coughing and difficulty in breathing) rather than binary variables, and then the proposed method could even be used for screening suspected cases. Second, with such data it would also be possible to model the likelihood of a pathogen in equation (1) not by arbitrarily measuring the distance but by using classification models using regression or more sophisticated machine learning approaches. Third, the erroneous input of incorrect information may be a challenge in real time analyses, although this did not appear to be an issue during the course of our analysis of the outbreak in Wuhan. However, it must be considered that the veracity of the source of information for such an analysis could have an impact on the resulting probability calculations. Fourth, the estimated probability that an outbreak is driven by a novel pathogen might be slightly over- or underestimated due to limited information about the mode of transmission and small numbers of observed cases. Of note, we believe that without 100% specificity of bacterial pathogens linked to the ongoing outbreak, excluding bacterial pathogens as candidate cannot be ensured, while the chance that the current outbreak is due to bacterial may be less suspected over time with partial clinical evidence. Nevertheless, the large number of characteristics that could be considered for the outbreak in Wuhan suggests that estimation was not beset in this study. Finally, we had to restrict ourselves to assume that the priori probability of all outbreak (***q***_***i***_) is identical. However, since the priori probability of observing the outbreak driven by a Disease X is completely unknown, we believe that this assumption can be plausible in this practice.

## CONCLUSIONS

Despite the future improvements to our statistical modelling framework that are required, this short study has demonstrated clearly that the ongoing outbreak of pneumonia cases in Wuhan is consistent with causation by a novel pathogen, “Disease X.” Analyses of the type conducted in this study can greatly support virological and genetic efforts to characterize the causal agent of this and future outbreaks, with the benefit that such analyses can be carried out extremely quickly.

## Data Availability

Supplemetal tables are available as attached to this submission.

## Author’s contributions

Sung-mok Jung: Data collection, formal analysis, model formulation, writing. Ryo Kinoshita: Data collection, formal analysis, visualization, writing. Robin N. Thompson: Data collection, model formulation, investigation, writing. Katsuma Hayashi: Data collection, visualization, writing. Natalie M. Linton: Data collection, model formulation, writing. Andrei R. Akhmetzhanov: Data collection, model formulation, writing. Yichi Yang: Data collection, writing. Hiroshi Nishiura: Conceptualization, model formulation, supervision, fund raising, validation, writing.

## Acknowledgements

The authors thank Dr Rebecca Spriggs for help devising the statistical approach. R.N.T. would like to thank Christ Church (Oxford) for funding via a Junior Research Fellowship. H.N. received funding from the Japan Agency for Medical Research and Development (AMED) [grant number: JP18fk0108050]; the Japan Society for the Promotion of Science (JSPS) KAKENHI [grant numbers, H.N.: 17H04701, 17H05808, 18H04895 and 19H01074; R.K.: 18J21587], the Inamori Foundation, and the Japan Science and Technology Agency (JST) CREST program [grant number: JPMJCR1413].

## Notes

### Competing Interest Statement

The authors have declared no competing interest.

